# FORGE: Functionally guided OCT Representation for Glaucoma Endophenotyping

**DOI:** 10.64898/2026.05.08.26352729

**Authors:** Mousa Moradi, Liyin Chen, Yan Zhao, Sayuri Sekimitsu, Niloufar Bineshfar, Janey L Wiggs, Mohammad Eslami, Mengyu Wang, Tobias Elze, Ayellet V. Segrè, Nazlee Zebardast

**Author notes:** These authors contributed equally to this work (Mousa Moradi, Liyin Chen). Corresponding author: Nazlee Zebardast.

## Abstract

Glaucoma endophenotyping is limited by disease heterogeneity and single-modality structural imaging. We introduce FORGE (Functionally guided OCT Representation for Glaucoma Endophenotyping), a cross-modal contrastive framework that uses visual field (VF) signals as privileged supervision during training, enabling OCT-only inference through a learned null embedding. In 5,372 paired examinations at Mass Eye and Ear, FORGE resolved 15 macular RNFL endophenotypes that fuzzy c-means (FCM) clustering partitioned into 9 clinically distinct phenotypic clusters, improving clustering quality over the OCT-only baseline by 22%. Transferred without refitting to 74,077 UK Biobank (UKB) and 3,119 Mass General Brigham Biobank images, they reproduced their defining RNFL-thickness signatures (Lin’s concordance 0.74). Treating each of the 15 continuous endophenotypes as a quantitative trait, genome-wide association analysis in 39,381 UKB European participants identified 53 LD-independent loci, 23 unreported for glaucoma or intraocular pressure, replicating in 711 external participants (sign concordance 75%). Endophenotypes learned with VF-guidance had increased genetic discovery power as compared with an identically trained ablation (median heritability Z 6.15 versus 4.76) and resolved retinal ganglion-cell enrichment.

## Introduction

Glaucoma (GL), the leading cause of irreversible blindness globally, is a heterogeneous neurodegenerative disease of the optic nerve ^1,2^. Clinical assessment relies on optical coherence tomography (OCT) to quantify retinal nerve fiber layer (RNFL) thickness and visual field (VF) testing to measure functional loss. Despite decades of research, substantial inter-individual variability persists in structural damage, functional impairment, progression rates, and therapeutic response ^3^, motivating clinically and biologically meaningful phenotyping beyond coarse clinical classification ^4^.

Robust phenotyping is essential for genome-wide association studies (GWASs) and imaging-genetic analyses ^5,6^. Poorly defined phenotypes dilute genetic signals, whereas representation learning captures biologically relevant variation beyond conventional summary measures ^7^. Most glaucoma (GL) imaging-genetic analyses rely on low-dimensional features including RNFL thickness ^8,9^, or cup-to-disc ratios ^10,11^, which fail to capture the full spectrum of glaucomatous damage that multimodal OCT-VF frameworks better integrate ^12^.

GL exhibits a stage-dependent dissociation and eventual coupling between retinal ganglion cell loss and functional impairment ^13^. OCT detects early structural damage, while standard automated perimetry captures functional loss patterns that often lag neural degeneration ^12^. Joint OCT-VF interpretation yields a more complete disease representation ^14^, reflecting the intrinsically entangled biological relationship between structure and function in this disease. Yet most computational phenotyping approaches remain single-modality, limiting clinical and biological characterization ^15^.

Most OCT and VF data remain unlabeled due to annotation cost and scalability constraints ^16^, necessitating self-supervised learning for medical image representation ^17^. Population-scale cohorts such as UK Biobank (UKBB) lack functional data ^18^, requiring methods utilizing multimodal training for structural-only inference. Contrastive learning methods such as Momentum Contrast (MoCo) learn representations by aligning related samples while separating distinct cases ^7,19^ and demonstrating efficacy in retinal imaging and imaging-genetics ^20,21^. However, existing approaches rely on single-modality approaches, require fully paired data ^22^, or obscure phenotypic diversity ^23^. These limitations are particularly problematic in GL, which comprises multiple endophenotypes with distinct structural and genetic architectures ^24,25^.

Recent progress in retina representation learning sharpens both the opportunity and the gap. Self-supervised objectives such as SimCLR and MoCo learn transferable embeddings without labels ^17,19^, and retinal foundation models trained on millions of images now generalize across ophthalmic and systemic tasks ^20^. In genetics, image-derived deep phenotypes have expanded discovery for optic-nerve-head morphology and other fundus traits beyond what handcrafted measures achieve ^6,7^. In parallel, large consortia have mapped the common-variant architecture of primary open-angle glaucoma and intraocular pressure through multitrait and cross-ancestry meta-analyses ^26,27^. establishing the reference against which any imaging-derived endophenotype must prove that it adds to, rather than merely recapitulates, known biology. Our own prior OCT-only endophenotyping showed that unsupervised structural representations already yield novel glaucoma loci ^28^; the unresolved question is whether functional supervision can be injected into such representations without sacrificing the structure-only deployment that population-scale genetics demands.

This is the central tension of computational glaucoma phenotyping: the most informative functional signal is precisely the one missing from the cohorts where phenotyping is most valuable, which has kept structure and function siloed. We resolve it by treating the visual field as privileged information ^29^, a signal that shapes the model during training and is then made dispensable at inference through a learned null embedding. The outcome is a single OCT encoder that carries functional knowledge yet runs on structure alone, producing endophenotypes that are simultaneously more coherent morphologically and more genetically informative than structure-only alternatives. Because the underlying principle, letting a sensitive but costly modality teach a scalable one, is not specific to glaucoma, functionally guided representation learning offers a general route from small, richly-phenotyped datasets to population-scale discovery.

To address these challenges, we introduce FORGE (Functionally guided OCT Representation for Glaucoma Endophenotyping), a cross-modal privileged-learning framework that integrates functionally guided contrastive learning with distribution-aware latent modeling to resolve mRNFL endophenotypes (Fig. 1). During training, FORGE uses the visual field as privileged functional supervision (formalized within the learning-using-privileged-information framework) to shape a shared β-VAE OCT representation, while a learned null embedding renders the VF dispensable at inference, so that endophenotypes can be assigned from structural imaging alone (Fig. 1A). We first derived VF-guided mRNFL representations in 5,372 paired clinical glaucoma examinations in Mass Eye and Ear (MEE), yielding 15-dimensional fuzzy c-means (FCM) and 20-dimensional Gaussian mixture modeling (GMM) latent endophenotypes that grouped participants into 9 and 13 phenotypic clusters, respectively (Fig. 1A); latent- and pixel-level analyses confirmed that VF guidance produced more coherent, better-separated phenotypes than the OCT-only baseline (Fig. 1B). We established the clinical validity of these endophenotypes independently of genetics, showing that they are eye-specific, associated with glaucoma and related ocular traits, and add discrimination beyond established clinical and polygenic predictors. We then transferred the frozen encoder, scaler, variational autoencoder (VAE), and cluster models, without refitting, to 47,252 UKBB and 1,453 MGBB participants, and confirmed that the endophenotype structure reproduced across these previously unseen cohorts (Fig. 1C). Finally, treating the reproduced endophenotypes as quantitative traits, we conducted genome-wide association analyses to test whether they are heritable and to identify the common-variant architecture underlying VF-guided mRNFL phenotypes, using UKBB for discovery and MGBB for replication, followed by post-GWAS characterization (Fig. 1C). Genetic analyses were conducted on the continuous latent axes rather than on discrete cluster membership. Cluster labels discard the graded within-cluster variation the latent space encodes and partition the cohort into unbalanced categorical groups, reducing power relative to a quantitative trait, whereas latent coordinates retain the full phenotype and are individually heritable. This follows established precedent for treating learned low-dimensional embeddings, rather than categorical assignments, as quantitative traits for genome-wide discovery ^30,31^.

**Fig. 1.**
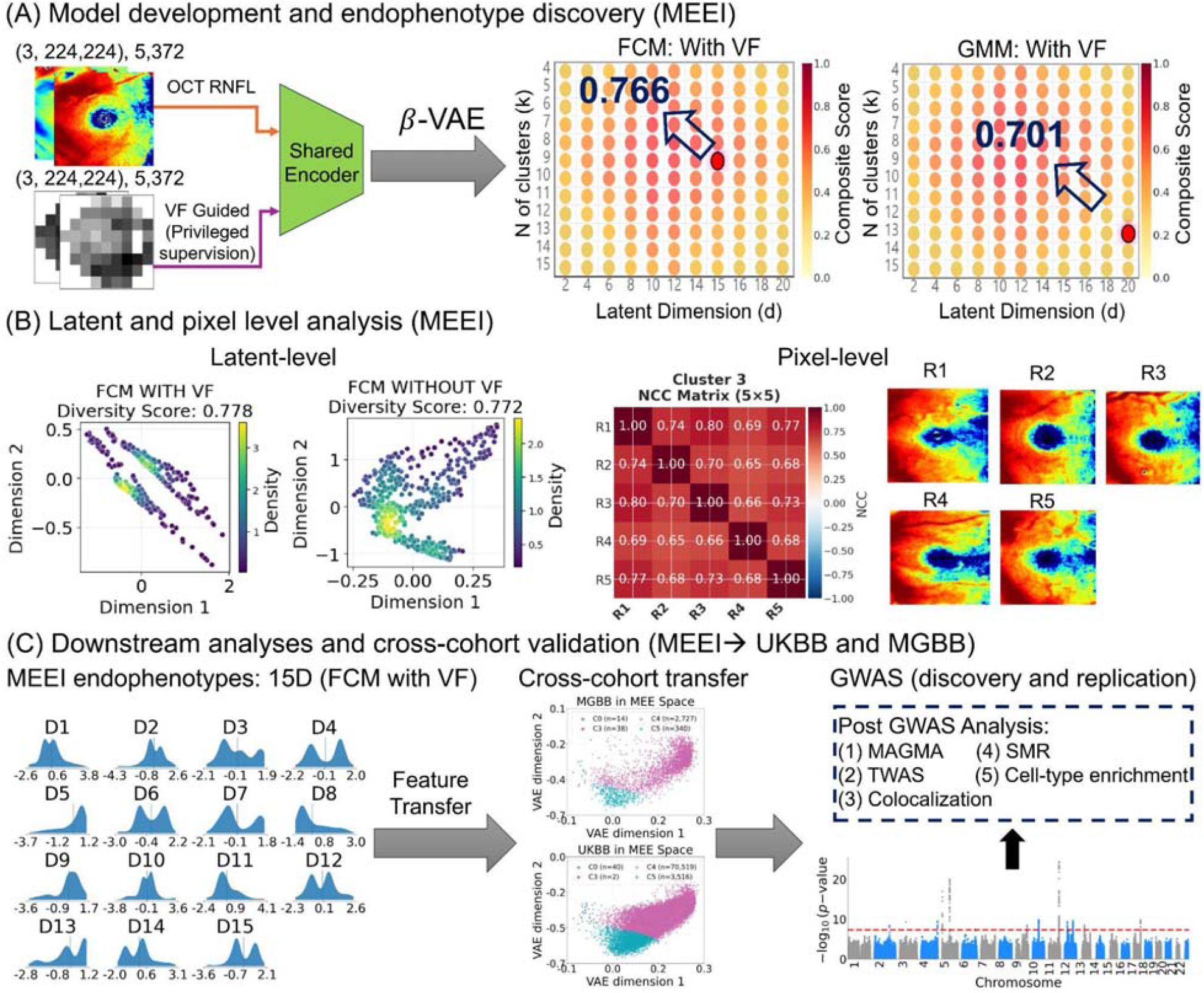
FORGE framework for glaucoma endophenotyping. (A) Paired OCT RNFL and VF images are encoded by a shared encoder trained with privileged VF supervision; a β-VAE sweep over latent dimension and cluster number (FCM and GMM, with VF) selects the model configuration by composite score. Each grid sweeps latent dimension (d, horizontal) against cluster number (k, vertical) and colors every configuration by its composite clustering score. In each grid the red marker is the selected configuration, and the open arrow is a callout linking it to its printed score (FCM, d = 15, k = 9, 0.766; GMM, d = 20, k = 13, 0.701). (B) Latent- and pixel-level analysis: diversity-scored latent embeddings (with vs. without VF) and per-cluster NCC matrices with representative RNFL maps. (C) Learned endophenotypes transfer cross-cohort to UK Biobank and Mass General Brigham Biobank for genome-wide discovery and replication, followed by post-GWAS analyses; OCT-only inference is enabled by replacing the VF input with a learned null embedding.

## Results

### Patient Demographics and Clinical Characteristics

Patient demographics and clinical characteristics for the development (MEEI) and external evaluation (UKBB and MGBB) cohorts are summarized in Table 1. Both external cohorts were younger than the development cohort (UKBB 57.1 ± 15.0 years; MGBB 59.1 ± 13.5 years vs MEE 69.0 ± 16.4 years; *P* < 0.001) and more racially homogeneous (89.0% and 74.9% White, respectively, vs 59.0%; *P* < 0.001), with MGBB intermediate between UKBB and MEE. Sex distribution differed modestly across cohorts (58.3%, 53.0%, and 58.9% female in MEE, UKBB, and MGBB; *P* < 0.001). Baseline mRNFL was thicker in both external cohorts than in the development cohort (UKBB 40.35 µm, +7.93 µm; MGBB 36.59 µm vs 32.42 µm; *P* < 0.001), whereas baseline IOP was comparable across all three cohorts (15.88 ± 5.00, 15.80 ± 3.97, and 15.49 ± 3.61 mmHg; *P* = 0.020, a difference of less than 0.4 mmHg that is unlikely to be clinically meaningful). Visual field data were available for MEE and MGBB only; baseline MD indicated substantially less advanced functional damage in MGBB (−3.85 ± 5.69 dB) than in MEE (−9.58 ± 7.43 dB; *P* < 0.001), consistent with a population-based biobank sampling rather than a tertiary glaucoma referral population. Together, the younger age, thicker mRNFL, and milder functional loss of both external cohorts frame the external evaluation as a test of generalization to less advanced and less selected disease.

**Table 1.** Patient demographics and clinical characteristics of internal and external datasets. Age, baselines mRNFL, MD, and IOP are presented as Mean ± SD. IOP= Intra ocular pressure.

| Characteristics | MEE Dataset<br>(*N=1,845) | UKBB Dataset<br>(N=47,252) | MGBB Dataset<br>(N=1,020) | P value |
| --- | --- | --- | --- | --- |
| Total number of scans | 5,372 | 74,077 | 3,119 |  |
| Age (years) | 69.0 ± 16.4 | 57.10 ± 15.0 | 59.1 ± 13.5 | < 0.001 |
| Gender, n (%) |  |  |  | < 0.001 |
| Female | 1660 (58.3) | 25,044 (53) | 601(58.9) |  |
| Race, n (%) |  |  |  | < 0.001 |
| Asian | 443 (24) | 3,308 (7) | 164(16.1) |  |
| Black or African American | 313 (17) | 1,890 (4) | 92(9) |  |
| White or Caucasian | 1,089 (59) | 42,054 (89) | 764(74.9) |  |
| Baseline mRNFL ($\mu\text{m}$) | 32.42 ± 4.48 | 40.35 ± 24.70 | 36.59 ± 9.29 | < 0.001 |
| Baseline MD (dB) | -9.58 ± 7.43 | NA | -3.85 ± 5.69 | < 0.001 |
| Baseline IOP (mmHg) | 15.88 ± 5.00 | 15.80 ± 3.97 | 15.49 ± 3.61 | 0.020 |

\*N= Number of patients. NA: Not Applicable (UKBB has no VF data).

### VF-guided phenotyping in the development cohort

On the MEE dataset (n = 538), VF-guided mRNFL representations achieved the strongest clustering performance under both FCM (15-dimensional latent space, m= 1.5, 9 clusters) and GMM (20-dimensional latent space, full covariance, 13 clusters), compared with the corresponding non-VF models (FCM: 20-dimensional latent space, 12 clusters; GMM: 20-dimensional latent space, 11 clusters; Fig. 2A, B). To isolate the contribution of functional guidance, the non-VF condition replaced the VF input with the learned null embedding while retaining the same architecture and training procedure, such that the two conditions differed only in the availability of VF information. VF guidance increased the composite clustering score by 22.0% for FCM (0.7661 versus 0.6282) and 11.3% for GMM (0.7008 versus 0.6297). This advantage was observed across all four-component metrics (the Silhouette score, normalized Calinski-Harabasz score, inverted Davies-Bouldin score, and diversity score) indicating that the improvement was not driven by a single clustering index (Fig. 2C; Table 2). Consistent with these findings, the latent density distributions showed more distinct and broadly distributed modes under VF guidance for both FCM and GMM (Supplementary Figs. 1 and 2, respectively). VF-guided representations also exhibited greater peak-location diversity than their non-VF counterparts for FCM (0.7655 versus 0.6547) and GMM (0.4969 versus 0.3874), further indicating that functional guidance expanded the distribution of phenotypic patterns across the latent space. At the pixel level, normalized cross-correlation analysis supported substantial within-cluster morphological coherence, with FCM yielding a higher mean NCC than GMM (0.4995⍰±⍰0.1958 versus 0.4318⍰±⍰0.1860). Representative patterns and within-cluster NCC matrices for the MEE clusters are shown in Supplementary Fig. 3 and 4 (with VF-guided) and Supplementary Fig. 5 and 6 (without VF-guided).

**Table 2.** Ablation Study. Effect of VF Guidance on mRNFL Phenotyping (MEE). Calinski-Harabasz scores are Normalized between 0-1. Davies-Bouldin scores were inverted so that higher values indicate better performance. Number of samples is the held-out test partition. W/O=Without.

| Metrics | Method |  |  |  |
| --- | --- | --- | --- | --- |
|  | FCM |  | GMM |  |
|  | With VF | W/O VF | With VF | W/O VF |
| **Number of samples** | 538 | 538 | 538 | 538 |
| **Latent Dimension** | 15D | 20D | 20D | 20D |
| **Number of clusters** | 9 | 12 | 13 | 11 |
| **Silhouette score** | 0.6042 | 0.4237 | 0.4801 | 0.4249 |
| **Calinski-Harabasz score** | 0.7450 | 0.3938 | 0.6196 | 0.3989 |
| **Inverted Davies-Bouldin score** | 0.9368 | 0.9218 | 0.9308 | 0.9209 |
| **Mean KL divergence** | 0.2018 | 0.1585 | 0.1809 | 0.1477 |
| **Mean entropy** | 3.3185 | 3.6802 | 3.2137 | 3.4983 |
| **Peak location diversity** | 0.7655 | 0.6547 | 0.4969 | 0.3874 |
| **Diversity score** | 0.7783 | 0.7725 | 0.7705 | 0.7644 |
| **Composite score** | 0.7661 | 0.6282 | 0.7008 | 0.6297 |

**Fig. 2.**
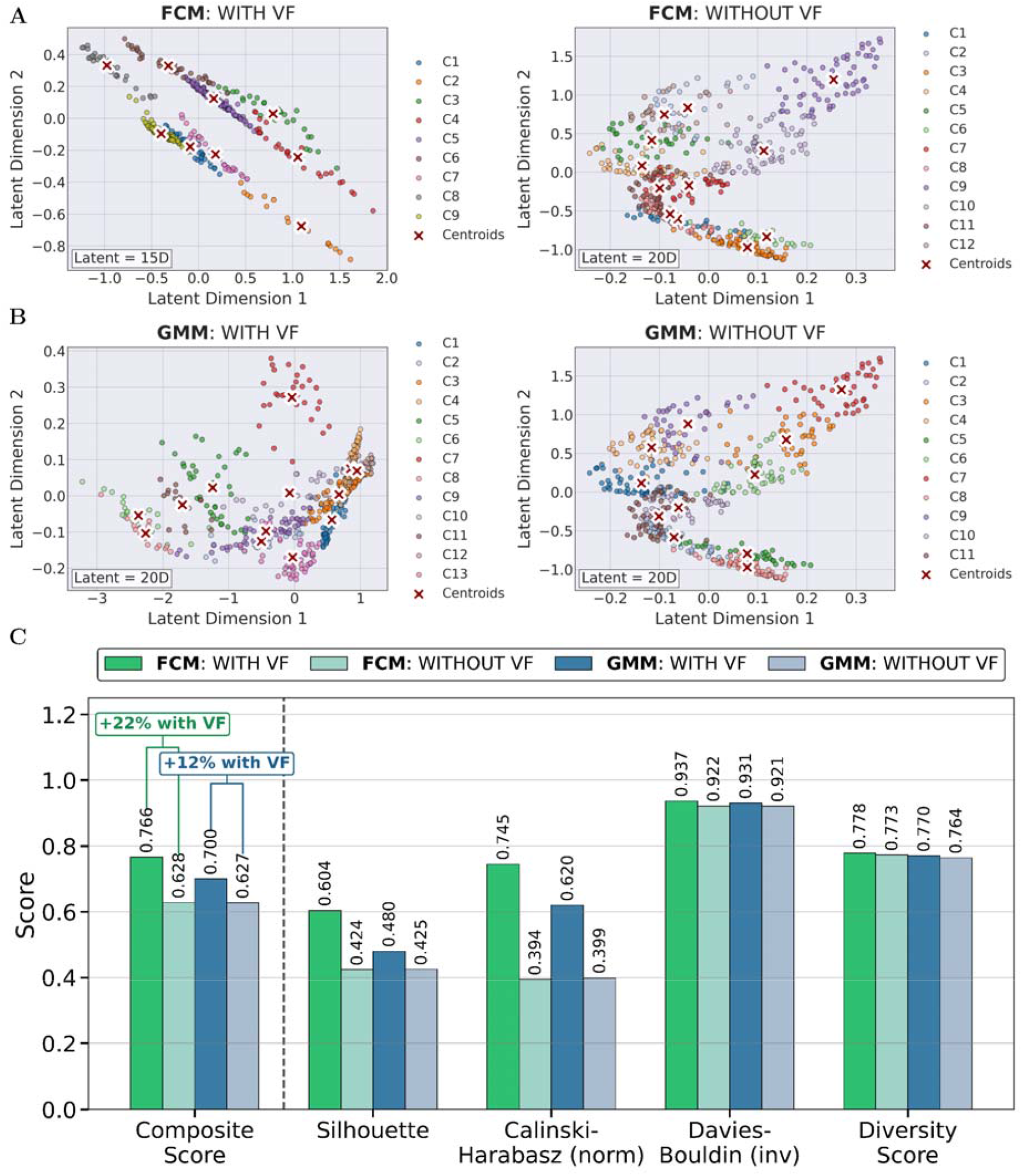
Latent space visualization and clustering quality metrics for FCM and GMM with and without VF data. (A, B) t-SNE projections of VAE latent representations showing cluster assignments for FCM (9 clusters, 15D) and GMM (13 clusters, 20D), with VF integration (left) improving cluster separation compared to OCT-only (right), (C) Quantitative performance metrics.

### Cross-cohort reproducibility of endophenotypes

To test whether the learned MEE representations generalize, the frozen encoder, scaler, VAE, and cluster models were transferred without refitting to two evaluation cohorts: UKBB (discovery) and MGBB (replication). Projecting each cohort into the MEE latent space recovered the FCM and GMM cluster structure in both UKBB (Supplementary Figs. 7 and 8) and MGBB (Supplementary Figs. 9 and 10). The correlation structure of the latent dimensions was comparable across both cohorts (Fig. 3): in UKBB, off-diagonal correlations averaged |*r*| = 0.472 (max 0.968; 50 dimension-pairs with |*r*| > 0.5) for the 15 FCM dimensions and |*r*| = 0.527 (max 0.991; 113 pairs) for the 20 GMM dimensions, with closely matching values in MGBB (FCM |*r*| = 0.404, max 0.944; GMM |*r*| = 0.585, max 0.994). The dimensions were therefore correlated rather than independent, the effective number of independent dimensions in UKBB was 5.0 of 15 (FCM) and 6.0 of 20 (GMM). We retained all 15 FCM axes as the units of genetic analysis and used Meff only to calibrate the significance threshold, following standard practice for testing correlated traits ^32^, rather than to reduce the traits themselves. Unlike the original axes, principal components cannot be assigned to a cluster or a reconstructed RNFL pattern, and replacing them would decouple the genetic analysis from the clustering, clinical-validity, and cross-cohort characterization performed on the same 15 axes throughout this study.

**Fig. 3.**
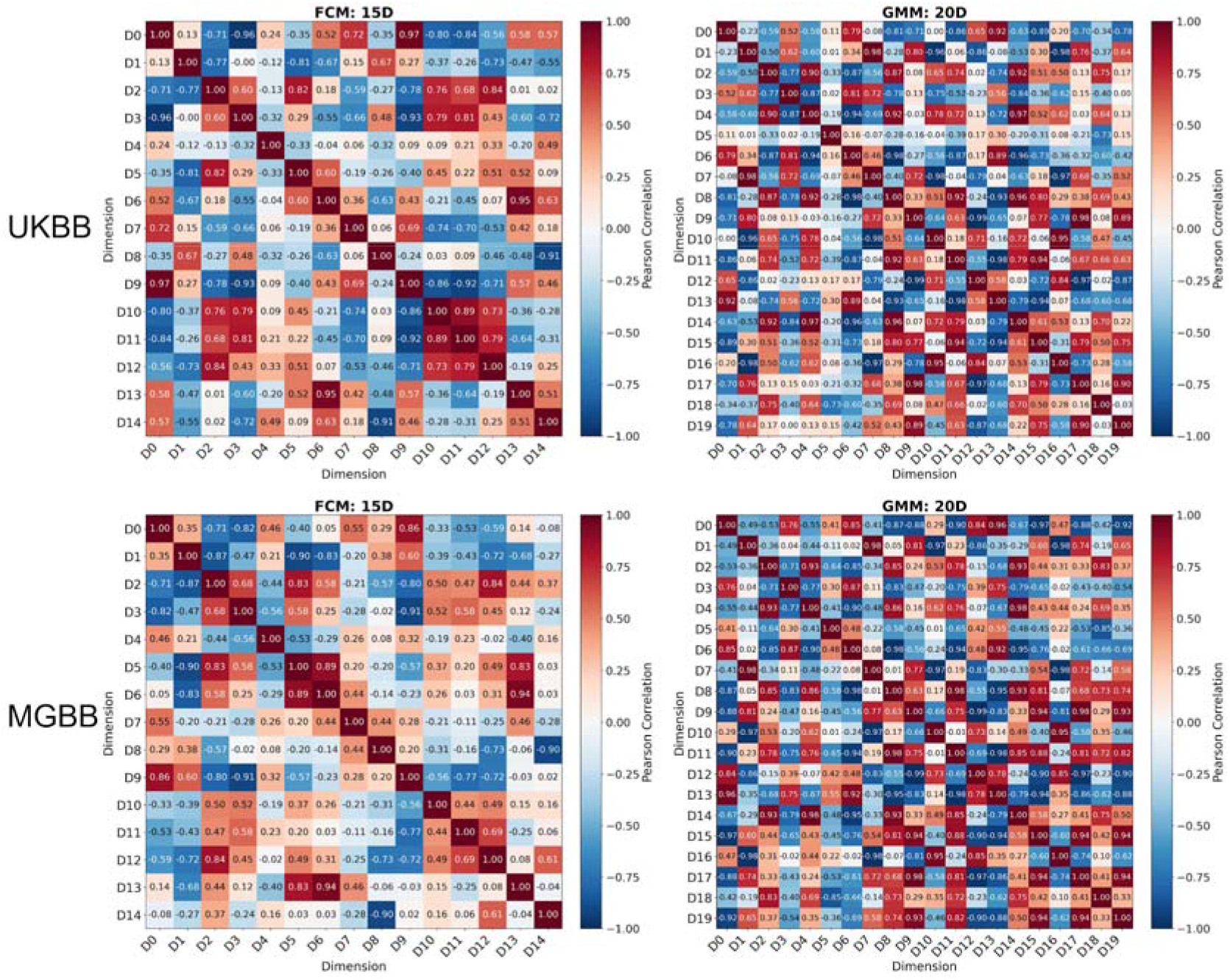
Latent-dimension correlation structure in the external cohorts. Pearson correlation matrices among the FCM (15D) and GMM (20D) latent dimensions in UK Biobank (top) and MGBB (bottom), computed on per-participant embeddings from frozen feature transfer. Consistent structure across cohorts indicates preserved representational geometry; the correlated dimensions reduce the effective number of independent traits (UKBB: 5.0/15 FCM, 6.0/20 GMM)

### Cross-cohort replication of clinical feature contributions to endophenotype assignment

To test whether the MEE-derived phenotypic structure itself reproduces externally, per-cluster signatures were reconstructed in each cohort, and their agreement was assessed against the identity line (Fig. 3). Within-cluster morphological coherence was preserved at population scale, though lower than internally: mean NCC was 0.304 (FCM) and 0.379 (GMM) across the populated UKBB clusters (*n* = 74,077 scans) and 0.295 (FCM) and 0.365 (GMM) in MGBB (*n* = 3,119 scans; Fig. 4A, B). Representative eyes and within-cluster NCC matrices for each cohort are shown in Supplementary Figs. 11 and 12 (MGBB, FCM and GMM) and Supplementary Figs. 13 and 14 (UKBB, FCM and GMM). Comparing matched clusters between the two external cohorts (*n* = 7), the defining structural signature, mean RNFL thickness, was highly concordant (Lin’s concordance correlation coefficient [CCC] = 0.742, Pearson *r* = 0.863, *P* = 0.012, bias = −1.3 µm; Fig. 4D), indicating that clusters occupy reproducible positions along the structural axis the model was trained to capture. In contrast, per-cluster mean IOP showed no concordance (CCC = −0.072, *r* = −0.160, *P* = 0.73; Fig. 4E), and the absolute magnitude of within-cluster coherence agreed only weakly (average NCC: CCC = 0.398, *r* = 0.419, *P* = 0.35; Fig. 4C), consistent with IOP and coherence magnitude being largely independent of the RNFL-structural phenotype and more sensitive to cohort-specific acquisition.

**Fig. 4.**
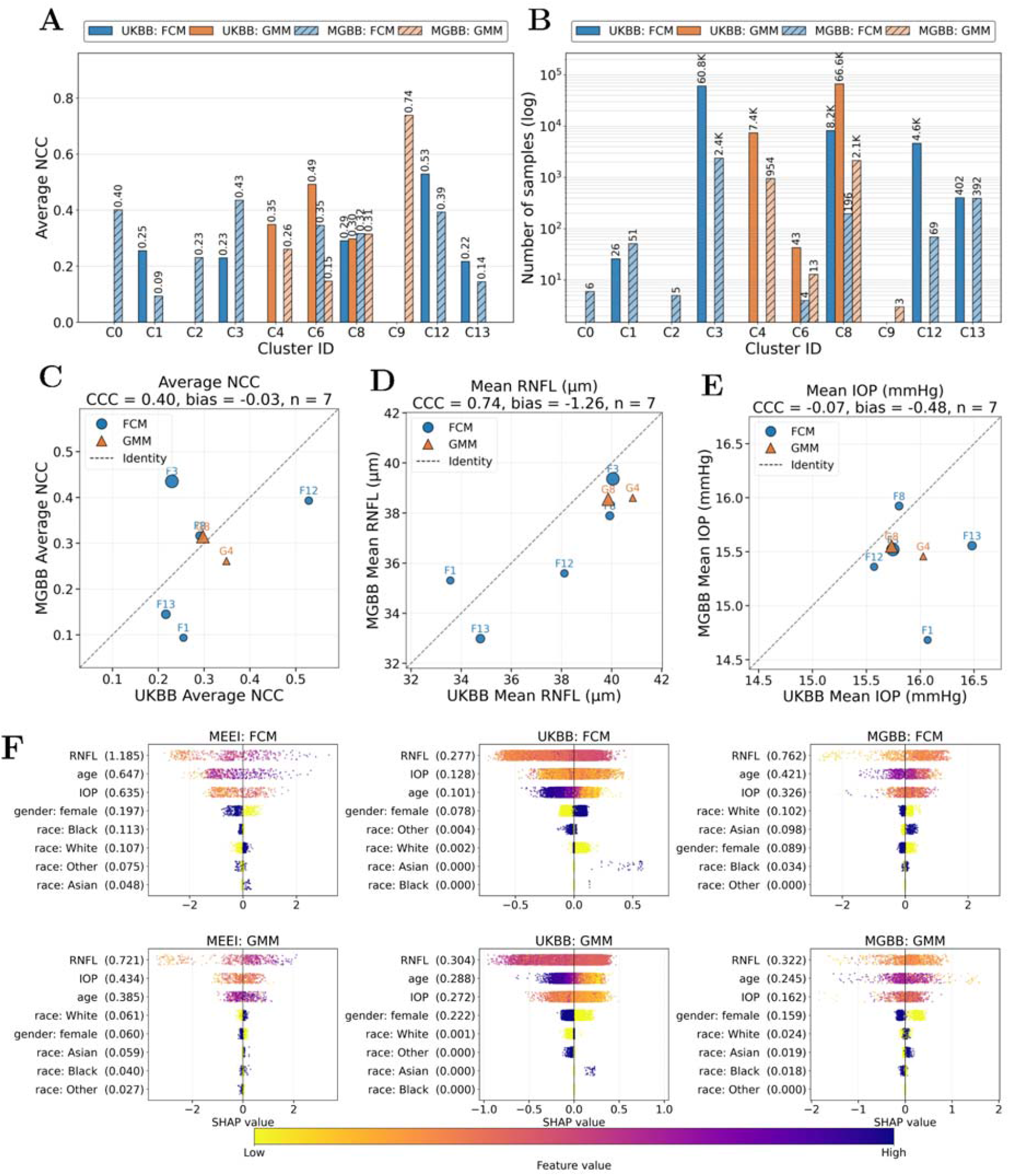
(A-E) Reproducibility of endophenotype signatures across external cohorts. (A, B) Per-cluster average NCC and sample sizes for UKBB and MGBB (FCM and GMM). (C-E) Cluster-level agreement between UKBB and MGBB against the identity line for average NCC, mean RNFL thickness, and mean IOP (Lin’s CCC and Pearson *r* annotated; *n* = 7 matched clusters), showing high concordance for RNFL thickness but not IOP. (F) Cross-cohort SHAP profiles of clinical features associated with endophenotype assignment. SHAP analysis of gradient-boosted classifiers predicting FCM (top) and GMM (bottom) cluster membership from RNFL, IOP, age, sex, and race in MEE, UKBB, and MGBB. Features are ordered by global mean absolute SHAP importance, with values shown in parentheses. Each point represents an individual participant; horizontal position indicates the signed SHAP value and color denotes the relative feature value from low to high.

To determine whether external reproducibility extended beyond latent-space geometry and cluster-level structural signatures to the clinically interpretable features associated with endophenotype assignment, we trained separate gradient-boosted classifiers in MEE, UKBB, and MGBB to predict FCM or GMM cluster membership from RNFL thickness, IOP, age, sex, and race, and quantified feature contributions using SHAP (Fig. 4F). The FCM analyses included 369 tested MEE participants (RNFL available in 431; IOP in 418), 47,252 UKBB participants (RNFL, 37,355; IOP, 41,490), and 1,020 MGBB participants (RNFL, 985; IOP, 924). The corresponding GMM analyses included 369 MEE participants (RNFL, 344; IOP, 331), 47,252 UKBB participants (RNFL, 37,366; IOP, 41,503), and 1,020 MGBB participants (RNFL, 1,018; IOP, 956).

RNFL had the highest mean absolute SHAP value in all six cohort-clustering combinations. For FCM, mean absolute SHAP values for RNFL were 1.185, 0.277, and 0.762 in MEE, UKBB, and MGBB, respectively; corresponding values were 0.647, 0.101, and 0.421 for age and 0.635, 0.128, and 0.326 for IOP. For GMM, mean absolute SHAP values for RNFL were 0.721, 0.304, and 0.322; values for age were 0.385, 0.288, and 0.245, and those for IOP were 0.434, 0.272, and 0.162 in MEE, UKBB, and MGBB, respectively. RNFL, age, and IOP comprised the three highest-ranked features in every cohort under both clustering approaches. Female sex had mean absolute SHAP values of 0.197, 0.078, and 0.089 for FCM and 0.060, 0.222, and 0.159 for GMM, while individual race-indicator contributions ranged from 0 to 0.113 for FCM and from 0 to 0.061 for GMM.

### Endophenotypes derived from FCM were chosen for further validation

Endophenotypes derived from FCM were used for the clinical-validity and genetic analyses that follow. FCM outperformed GMM on all four components of the configuration-selection composite score (Silhouette, normalized Calinski-Harabasz, inverted Davies-Bouldin, and diversity score), yielding a higher composite score (0.766 versus 0.701), greater peak-location diversity (0.766 versus 0.497), and a larger improvement attributable to visual-field guidance (22.0% versus 11.3%). In an independent pixel-level check, FCM clusters also showed higher within-cluster morphological coherence (mean NCC 0.500 ± 0.196 versus 0.432 ± 0.186). We retained GMM as a robustness check on the phenotyping step: that two mechanistically distinct clustering algorithms agreed on the direction and presence of the visual-field effect indicates the effect is a property of the learned representation rather than of a particular clustering method. All results reported below refer to the 15 FCM endophenotypes

### The endophenotypes are eye-specific and strongly associated with glaucoma

Phenotypic validity was assessed against three pre-specified families of outcomes in UKBB (Fig. 5A, Supplementary Table 1) to establish that the endophenotypes act as retinal-specific traits. Each family asks a different question of them, namely, whether they reproduce on ocular traits independent of OCT; and whether they are specific to retinal traits.

**Fig. 5.** The VF-guided endophenotypes are eye-specific, offer incremental discrimination value, and provide increased genetic discovery power. (A) Association of the 15 axes with 16 outcomes in three pre-specified families. Left, odds ratio per s.d. for POAG (325 cases) on a log axis with 95% Wald intervals; filled circles mark FDR < 0.05 within outcome, which 14 of 15 axes reach. Right, signed −log10 P from the likelihood-ratio test, on a diverging scale clipped at ±20; asterisks mark FDR < 0.05 within outcome; the three blocks are ocular disease, ocular quantitative traits, and one systemic negative control per organ system. (B) Cross-validated AUROC for four cumulative models. Points are AUROC and lines are 95% intervals combining the DeLong standard error with the across-seed variance over five outer seeds. The right-hand column is what the 15 endophenotypes add over that row, with *** P < 0.001, ** P < 0.01 and * P < 0.05. C) The 105 endophenotype pairs, genetic correlation below the diagonal and phenotypic above, on one average-linkage ordering of 1 − |rg|. Each cell is a wedge whose filled fraction is |r| and whose colour is r. A grey cross marks a genetic estimate that does not clear Bonferroni across the 105 pairs; red shading marks a pair whose two estimates disagree in sign, shaded in both of its cells. D) Comparison of LD score regression heritability Z between VF-guided endophenotypes and endophenotypes derived without VF guidance. Bars are medians and the red line is the conventional reliability threshold Z = 4, which 15 of 15 visual-field-guided axes clear against 18 of 20 ablation axes. P is Mann-Whitney. E) Comparison of the number of loci, physical regions and novel loci between the two representations.

The first family asks whether they track disease of the retinal layers. Against primary open-angle glaucoma (POAG) defined by ICD codes, 14 of the 15 FCM endophenotypes were significant at Benjamini-Hochberg FDR < 0.05 and the largest effect reached an odds ratio of 2.17 per standard deviation. Similarly, against the broader any-glaucoma definition (ICD and self-reported), all 15 were significant, with the largest odds ratio being 1.84 at *P* = 9.0 × 10^−69^. Associations with the other ocular diseases were present but consistently weaker.

The second family asks whether the endophenotypes are associated with ocular traits that are independent of OCT measurements. Significant associations support that these endophenotypes capture ocular information beyond what OCT imaging provides. The results showed that 13 of the 15 endophenotypes significantly (FDR < 0.05) were associated with intraocular pressure (IOP) (11 of them at the study-wide threshold), 13 significantly associated with corneal resistance factor, and 8 for cup-disc ratio. The last family, acting as a negative control, checks seven systemic conditions with no plausible retinal-morphology pathway, one drawn from each organ system tested, where. Six of the 7 conditions did not show any significant signal (FDR < 0.05), supporting that the endophenotypes behave as retinal traits rather than as a marker of general ill health.

### The endophenotypes add glaucoma discrimination value

Discrimination was assessed by cross-validated ridge-penalized logistic regression across four cumulative models: 1) basic demographics (age and sex), 2) demographics plus clinical measurements (IOP and baseline RNFL thickness), 3) demographics, clinical measurements, and a representative POAG polygenic risk score ^27^, and 4) all of above plus the 15 endophenotypes. These models were fitted on the same 34,411 participants who have all the required measurements available.

The endophenotypes improved discrimination over every baseline tested. For POAG, cross-validated AUROC rose from 0.716 (95% CI 0.688 to 0.744) with age and sex alone, to 0.823 (0.797 to 0.850) once intraocular pressure and RNFL thickness were added, 0.838 (0.813 to 0.863) with the polygenic score, and 0.861 (0.838 to 0.884) with the 15 endophenotypes. Measured against each of those baselines in turn, the endophenotypes added +0.145 (0.117 to 0.174, *P* = 5.2 × 10^−23^), +0.038 (0.019 to 0.056, *P* = 7.0 × 10^−5^) and +0.023 (0.008 to 0.038, *P* = 2.8 × 10^−3^). The pattern was the same for any glaucoma, where AUROC rose from 0.691 (0.672 to 0.710) to 0.823 (0.807 to 0.840) across the same four models, with increments of +0.132 (0.114 to 0.151, *P* = 2.2 × 10^−44^), +0.022 (0.013 to 0.031, *P* = 1.8 × 10^−6^) and +0.014 (0.007 to 0.022, *P* = 2.6 × 10^−4^). Because mean macular RNFL thickness already sits in the baseline, the final increment is what the endophenotypes contribute over the same macular scans they are derived from, on top of pressure and polygenic risk (Fig. 5B, Supplementary Tables 2 band 3). In addition, we confirmed that the gain from the 15 endophenotypes was not due to model capacity by running a permutation control. We replaced the 15 endophenotypes with permuted copies of matched dimensionality, fit the model through the identical nested-CV ridge pipeline, and found this control yielded no improvement over model 3 (ΔAUROC = 0.836, 95% CI [0.811-0.861]), in contrast to the +0.023 gain from model 4.

### Genetic correlation structure mirrors the phenotypic one

GWAS was performed on the 15 FCM endophenotypes derived under VF-guidance and on the 20 axes derived by the same procedure without VF guidance, in 39,381 UK Biobank participants of European genetic ancestry. Participants had a mean age of 56.9 years (s.d. 8.0) and 54.1% were female. 9,050,596 autosomal variants were tested per endophenotype after a genome-wide call-rate filter applied before locus definition. Test-statistic inflation was modest and consistent across traits (λ_GC_ 1.025-1.064, median 1.051, univariate LD score regression intercepts of 0.996 to 1.023), indicating that there was no evidence of inflation driven by population stratification or cryptic relatedness.

Because these endophenotypes were defined phenotypically, their value as genetic traits rests on that phenotypic structure having a genetic counterpart. We tested this by comparing the phenotypic and genetic correlation matrices across all 105 pairs. The endophenotypes are strongly inter-correlated, with a median |*r*_g_| of 0.65, and the two matrices agree closely (Fig. 5C). Each pair carries a phenotypic correlation estimated in the genotyped participants and a genetic correlation estimated by cross-trait LD score regression. The uncentred correlation between the two matrices was 0.925 and 94 pairs shared the same sign.

For the eleven pairs whose two estimates disagree in sign, shaded in red in both cells of the pair in Fig. 5C, none had a genetic correlation that clears Bonferroni correction across the 105 pairs. In other words, no disagreement occurred where the genetic estimate is resolved. Among the 76 pairs whose genetic correlation was resolved, sign agreement is 76 of 76. Genetic correlations are overall the larger in magnitude, with a ratio of mean |*r*_g_| to mean |*r*_p_| of 1.28 (Supplementary Table 4).

### Visual-field guidance improves genetic discovery strength

Every endophenotype derived from the VF-guided method showed significant common-variant heritability by two estimators (LD score regression, observed-scale *h*^2^ 0.061-0.120, *Z* 4.98-7.80; GCTA-GREML 0.083-0.201; Supplementary Fig. 15, Supplementary Table 5). The two estimators were strongly concordant (Pearson *r* = 0.83).

Compared with the endophenotypes derived without VF guidance, the VF-guided ones offer more genetic discovery power. Median observed-scale *h*^2^ is 0.096 against 0.073 (*P* = 0.029), and median mean χ^2^ − 1 is 1.31-fold larger (*P* = 1.7 × 10^−3^). Median LD score regression *h*^2^ *Z* is 6.15 in VF-guided against 4.76 for no-VF (Mann-Whitney *P* = 9.0 × 10^−5^) (Fig. 5D, Supplementary Table 6). Restricting the ablation to its 15 most heritable axes leaves the difference intact (median *Z* 4.93, *P* = 9.1 × 10^−4^). In addition, the no-VF representations yielded 40 LD-independent loci in 32 physical regions (18 novel signals) which are fewer than the VF-guided comparator on every number (Fig. 5E, Supplementary Table 7).

### GWAS identifies 53 independent loci in 41 physical regions, 23 of them novel

Per-endophenotype LD clumping (*r*^2^ < 0.1, ±500 kb window, index *P* < 5 × 10^−8^) yielded 160 independent index variants. Pooling these across traits and re-clumping under identical parameters resolved them into 53 LD-independent loci, collapsing into 41 distinct physical regions (Fig. 6A, Supplementary Tables 8 and S9). Twenty-two of the 53 loci (42%) were associated with more than one endophenotype. Since the endophenotypes are intercorrelated, we applied dimensional reduction and identified an effective number of independent dimensions, *M*_eff,_ as 5 ^32^ (Supplementary Fig. 16, Supplementary Table 10). For the genetics analyses below, a Bonferroni multiplying testing correction of 15 is used as the primary, conservative threshold, and *M*_eff_ = 5 is used as the effective-dimensionality tier.

**Fig. 6.** Genetic discovery in UKBB and replication in MGBB. (A) The 15 GWAS overlaid on one axis. Variants below genome-wid significance are grey, alternately shaded by chromosome and thinned on a pixel grid; variants reaching P < 5 × 10^−8^ (red line) are drawn in full and coloured by the number of axes reaching that threshold at the same variant, from 1 to 14. Labels name the nearest protein-coding gene of each of the 41 regions, one per region. (B) UpSet of the 53 leads against the seven previously reported reference sources, each lead classified hierarchically as LEAD, PROXY, PROXIMITY or no match (Methods). Filled dots mark membership and the rule spans one intersection. Evidence hierarchy is preserved by stacking each bar by the strongest class its loci achieve, and the tinted degree-0 column at the far left is the 23 loci matching no source. A donut marks an exact lead-variant hit, which is the only class requiring no LD assumption. (C) Discovery against replication effect sizes, in standard deviations of the axis per copy of the effect allele. The two axes carry identical limits and an equal aspect ratio, so the dashed grey line is the line of identity. Grey whiskers are 95% Wald intervals on the replication estimate; the discovery intervals ar omitted for legibility. The orange line is the Deming errors-in-variables slope, and the shaded band is its locus-block bootstrap interval. The permutation P refers to sign concordance. (D) POAG association per standard deviation of each axis, with 95% Wald intervals on a log axis; filled circles mark FDR < 0.05 across the 15 axes and the dashed line at 1 is the null.

The 53 loci were compared against previously reported associations of POAG, IOP, and our previous OCT-only endophenotype GWAS. Twenty-six of the 53 loci are novel to POAG and IOP, and the number decreased to 23 when the loci from our previous OCT-only endophenotype GWAS was included as the reference. These 23 novel loci occupied 21 distinct physical regions (Fig. 6B, Supplementary Table 11). The novel set is identical under both proxy thresholds (*r*^2^ ≥ 0.2 and *r*^2^ ≥ 0.8). At the *M*_eff_ tier (*P* < 1.0 × 10^−8^), 37 loci yield 16 novel signals in 15 regions. The novel loci carry association strength comparable to the reported ones (median −log_10_ *P* 9.09 versus 9.78, Mann-Whitney *P* = 0.54), and the 41 regions partition cleanly, with no region containing both a novel and a previously reported lead.

Statistical fine-mapping of all 41 regions in their strongest trait returned 47 credible sets across 40 regions, with a median purity of 0.919 (Supplementary Table 12). Eight sets resolve to a single variant and 23 of 47 (48.9%) contain ten or fewer.

### The genetic architecture and the clinical association replicate in MGBB

We tested replication in 711 unrelated MGBB participants of European genetic ancestry, in whom genomic inflation was negligible (median λ = 1.005). The 105 testable lead variant-axis pairs comprised 53 unique variants in 26 independent 1 Mb loci (Supplementary Table 13). Because pairs within a locus are not independent (design effect 4.18, equivalent to 25.1 independent observations), we clustered all inference below on the locus.

Discovery and replication effects agreed in both direction and magnitude. Signs were concordant for 79 of the 105 pairs (75.2%, locus-block permutation P = 0.002), the two sets of estimates correlated at r = 0.60 (95% CI 0.30-0.72, locus-block bootstrap), and the Deming errors-in-variables slope was 0.74 (95% CI 0.39-1.05; Fig. 6C). Median per-variant power was 0.235 at one-sided α = 0.05, and no pair reached 80% power at any threshold. The results therefore support replication of genetic architecture but lack power for variant-level replication.

The axes also carried their clinical association into MGBB. Among the 435 unrelated participants with POAG status (280 cases and 155 controls), 14 of 15 axes were associated with POAG at FDR < 0.05, with the largest effect being 1.88 per SD (95% CI 1.45-2.43; Fig. 6D, Supplementary Table 14).

### Four converging lines of genetic evidence nominate 28 putative causal genes, three at novel loci

We applied four complementary analyses to prioritize genes. MAGMA gene-based association, which aggregates variant-level statistics across each gene and its flanking region, identified 36 genes at the primary threshold of 0.05/ (18,417 × 15) and 39 at the Meff tier. Every endophenotype had at least one significant gene (Supplementary Table 15). Transcriptome- and splicing-wide association was performed with S-PrediXcan and aggregated across 18 tissues using S-MultiXcan. Thirty genes reached the primary threshold and 36 reached the Meff tier, comprising 33 expression-associated genes and 18 splicing-associated genes, with 15 supported by both. Of these 36 genes, 22 were also significant in the MAGMA analysis (Supplementary Table 16).

Colocalization tested whether regional GWAS associations and cis-e/sQTL signals were consistent with a shared causal variant. The strongest endophenotype association in each region was tested against cis-e/sQTL data from 18 GTEx tissues and retina (EyeGEx, n=406), yielding 2,579 tests. The GTEx analysis covered 97.6% of gene-level tuples and all tuples in regions with a GWAS credible set (Supplementary Tables S17 and S18). Called signals were subsequently examined using locus-compare plots. Eight genes were supported: OAF (PP4 = 0.996 in retina and 0.989 in GTEx; Pearson r=+0.95), MIR31HG (PP4 = 0.990; r=+0.77), ADCY5 (0.985; +0.51), TSPAN11 (0.983 across 10 GTEx tissues and 0.981 in retina; +0.87), TSPAN10 (0.967; +0.81), CNKSR3 (0.953; +0.85), IFNE (0.939; +0.80) and ARHGEF7 (0.928; +0.93).

SMR with the accompanying HEIDI test provided an additional assessment of shared association versus linkage. Retina was the primary tissue, with twelve GTEx tissues used to assess cross-tissue support. Eleven genes passed the arm-specific SMR threshold and HEIDI P>0.05, (Supplementary Table 19).

A putative causal gene was defined as a gene supported by at least two of the four analyses (Fig. 7A). Twenty-eight genes met this criterion, nine of which were supported by at least three analyses. ARHGEF7, OAF and TSPAN10 were supported by all four. Three genes, ARHGEF7, CNKSR3 and ADCY5, which were genes novel to POAG, IOP, and our previous OCT-only endophenotype GWAS, were supported in retina by both colocalization (Fig. 7B) and SMR.

**Fig. 7.**
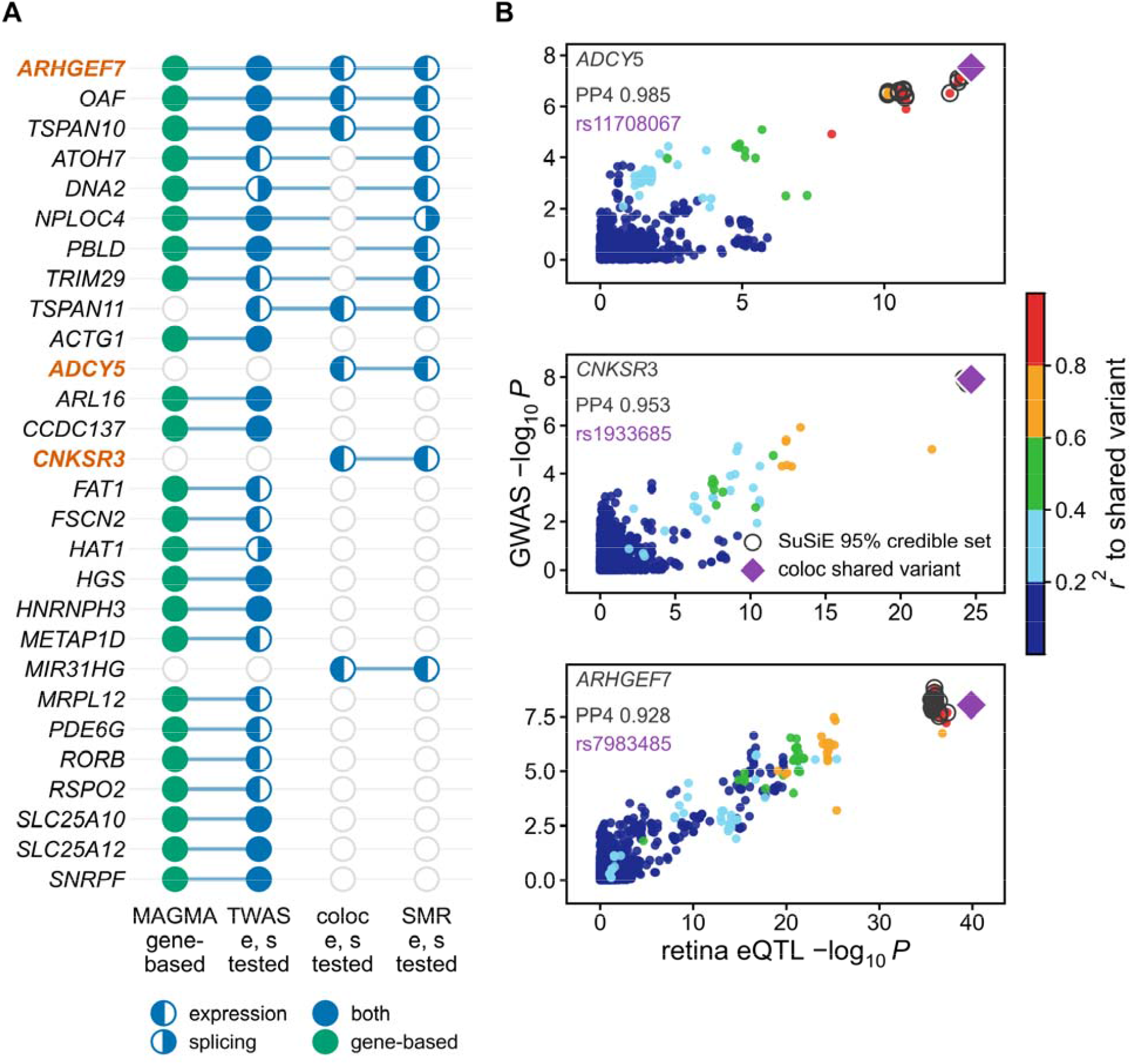
Four independent lines of evidence prioritized 28 genes, with three at novel loci. (A) Support from gene-based association, transcriptome- and splicing-wide association, colocalization and summary-data-based testing, for the 28 genes supported by at least two. A filled mark is a supported line and the connecting rule spans the supported set, so a full row reads as one bar. The three names colored, ADCY5, CNKSR3 and ARHGEF7, are the genes at novel loci. (B) Locus comparison at those three loci. Each point is a variant present in both scans, plotting retina cis-eQTL against GWAS −log10 P for the region’s strongest trait, colored by r2 to the shared variant. The vermillion diamond is the shared variant colocalization itself nominated. Open rings are members of that region’s 95% credible set from the GWAS fine-mapping, and in all three the nominated variant is a credible-set member, so the two methods agree on which signal is being explained. PP4 is the coloc statistic that represents the posterior probability of both traits sharing the same causal variant for a given locus.

### Visual-field guidance resolves the ganglion-cell component of the cell-type signal

We performed ECLIPSER to test whether genes mapped to genome-wide significant endophenotype loci were enriched for cell-type-specific expression relative to null GWAS loci. The analysis was run against four ocular single-cell references, namely, retina, macula, optic nerve head and anterior segment (Supplementary Table 20). Six tests reached a within-reference Benjamini-Hochberg-adjusted P < 0.05, all in the retina reference (Fig. 8A). Müller glia was the strongest, at a fold-enrichment of 3.73 (P = 0.010), followed by retinal pigment epithelium (3.01, P = 9.5 × 10^−3^) and astrocytes (2.94, P = 0.011). The concentration of signal in glia and pigment epithelium is consistent with previous POAG cell-type enrichment analyses ^33^. More importantly, our analysis also identified ganglion cells enrichment: ON midget (3.06, P = 8.9 × 10^−3^), OFF midget (2.95, P = 0.011) and other ganglion cells (2.93, P = 0.011).

**Fig. 8.**
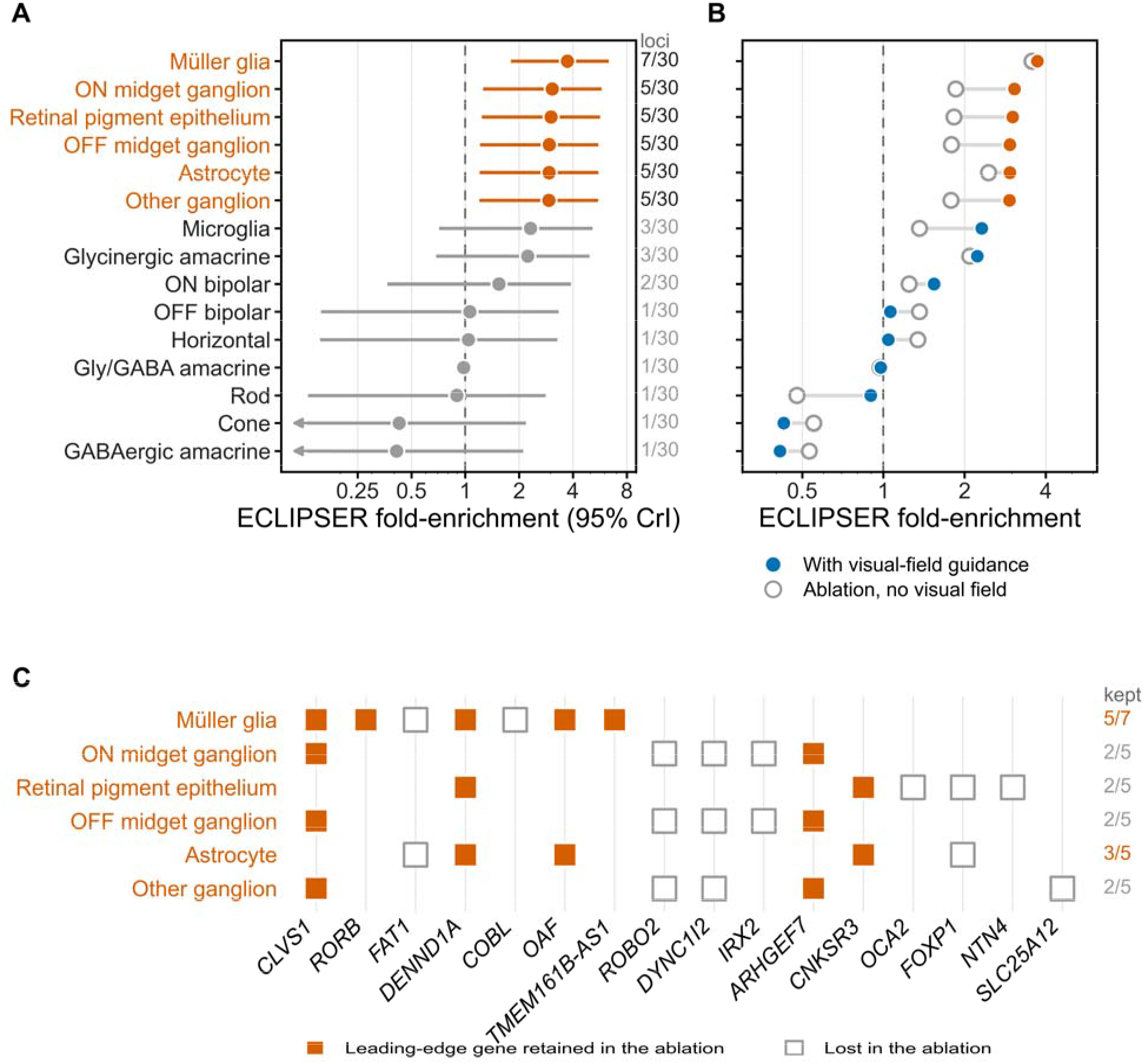
Locus-based cell-type enrichment localizes the common-variant signal to Müller glia, pigment epithelium and midget ganglion cells, and visual-field guidance is what resolves all but the glial component. (A) ECLIPSER fold-enrichment in the retin reference under visual-field guidance. Points are posterior median fold-enrichments and lines are 95% credible intervals on log axis; arrows mark intervals extending beyond the plotted axis. Vermillion marks the six cell types reaching Benjamini-Hochberg-adjusted P < 0.05 within the reference. The right-hand count gives the number of the 30 gene-mapped loci that ar leading-edge loci for each cell type. (B) The same 15 cell types under both representations, point estimates only. (C) The leading edge itself, for the six cell types resolved in (A). A filled square marks a leading-edge gene retained in the ablation and an open square one lost; the right-hand count is the number retained.

The same analysis was applied to the No-VF endophenotypes to assess the contribution of visual-field guidance. The ablation produced fewer gene-mapped loci in every reference: 23 versus 30 in retina, 22 versus 29 in optic nerve head, 19 versus 27 in anterior segment and 17 versus 25 in macula (Supplementary Fig. 17). No cell type reached the within-reference threshold in the ablation, compared with six under visual-field guidance. The attenuation varied across cell types (Fig. 8B). Müller-glia enrichment was largely preserved, with a fold-enrichment of 3.56 compared with 3.73 under visual-field guidance. Five of its seven leading-edge loci were retained, and Müller glia remained the leading cell type in the reference, although its adjusted P value increased to 0.057. Astrocytes retained three of five leading-edge loci, with a fold-enrichment of 2.45. Retinal pigment epithelium and each of the three ganglion-cell annotations lost three of five leading-edge loci (Fig. 8C), and their fold-enrichments fell by approximately two-fifths and became indistinguishable from the null background. These results indicated that visual-field guidance is what resolves the ganglion-cell signal.

## Discussion

FORGE was designed to test a single premise: that visual-field function, available only during training, can be distilled into OCT representations so that structural imaging alone remains functionally informed at inference. On the MEE dataset, VF guidance improved the composite clustering score by 22.0% for FCM and 11.3% for GMM, with concordant improvements across the Silhouette, normalized Calinski-Harabasz, inverted Davies-Bouldin, and diversity metrics. The latent density distributions likewise showed more distinct and broadly distributed modes under VF guidance for both clustering approaches (Supplementary Figs. 1 and 2), accompanied by greater peak-location diversity for FCM and GMM. Normalized cross-correlation further demonstrated substantial within-cluster morphological coherence, with higher mean NCC for FCM than for GMM. Together with the reproducibility of clinical feature contributions across cohorts and the recovery of established loci and candidate novel genetic associations, these observations support the premise that functional guidance identifies biologically meaningful axes of structural variation. The concordance across two mechanistically distinct clustering algorithms further suggests that these axes reflect properties of the learned representation rather than artifacts of a particular partitioning method. FCM was carried forward for all downstream clinical-validity and genetic analyses, as it outperformed GMM on every selection criterion and yielded higher within-cluster coherence; GMM was retained only as this cross-algorithm robustness check. In UK Biobank, the resulting 15 FCM endophenotypes were heritable, yielded 53 LD-independent loci, and were associated with POAG. The effect sizes of the loci replicated in direction and magnitude in MGBB, as did the clinical associations.

The SHAP analysis provided an additional level of validation by testing whether the same clinically interpretable variables contributed to subtype assignment across the development cohort and two independent external cohorts. RNFL was the dominant feature in all six cohort-clustering combinations, with mean absolute SHAP values ranging from 0.277 to 1.185 for FCM and from 0.304 to 0.721 for GMM. Age and IOP consistently ranked among the three most influential features, whereas sex and individual race indicators generally contributed less. This hierarchy was observed across MEE, UK Biobank, and Mass General Brigham Biobank despite substantial differences in cohort composition, disease severity, and sampling context. The repeated prominence of RNFL is consistent with the structural basis of the learned endophenotypes, whereas the contributions of IOP and age indicate that cluster membership captures clinically relevant variation beyond retinal thickness alone. Thus, external replication was not limited to latent geometry or mean cluster morphology; the clinical variables associated with subtype assignment were also broadly preserved across cohorts.

To prioritize effector genes, we required convergence across independent lines of evidence. Twenty-eight genes were supported by at least two of gene-based association, transcriptome- and splicing-wide association, colocalization, and MR, and ARHGEF7, OAF, and TSPAN10 were supported by all four. The 3 prioritized novel loci (ARHGEF7, CNKSR3, and ADCY5) were each supported in retina by both colocalization (PP4 0.928 to 0.985) and SMR, with the nominated variants inside the fine-mapping credible sets. The functions of these three genes suggest relevance to retinal neuronal integrity. ADCY5 couples G-protein-coupled receptor activation to cAMP production ^34^; CNKSR3 scaffolds membrane-associated signalling and epithelial ion-transport complexes ^35^; and ARHGEF7/β-PIX links Rho-family GTPases to actin remodelling, synaptic organization and neuronal axon formation ^36–38^. Their functions span membrane-proximal signal transduction to cytoskeletal and synaptic organization. The wider prioritized gene set reinforced several themes relevant to retinal biology, including cytoskeletal organization (ACTG1 and FSCN2) ^39,40^, membrane trafficking (TSPAN10 and HGS) ^41,42^, and retinal neuronal development (ATOH7 and RORB) ^43,44^. These annotations suggest that POAG susceptibility may arise from modest perturbations across processes that collectively sustain retinal cell identity, metabolic competence and neuronal connectivity, which lowers the threshold for retinal ganglion-cell dysfunction under glaucomatous stress.

Cell-type analyses localized the polygenic signal within the retina. Müller glia showed the strongest enrichment, followed by retinal pigment epithelium, astrocytes, and the ON midget, OFF midget, and other ganglion-cell populations. The glial component of this signal is concordant with the reported enrichment of POAG-colocalizing genes in retinal astrocytes and Müller glia ^33^, which supports the calibration of the pipeline. The enrichment of midget ganglion-cell populations at the discovered loci is the observation that aligns most directly with the defining neuronal pathology of glaucoma (i.e. the degeneration of retinal ganglion cells and their axons ^45^), and it emerged only when functional supervision shaped the phenotypes. This pattern suggests that VF guidance concentrates the representation on structural variation whose genetic regulation acts through the cells that die in the disease as well as through the glial and epithelial populations that support them.

Methodologically, the learned null embedding is central to the deployability of the framework. By learning an explicit representation of the unavailable modality, the multimodal encoder supports OCT-only inference without retraining or architectural modification. This design permits direct application to large imaging biobanks in which perimetry is unavailable for most participants. The approach places glaucoma endophenotyping within the learning-using-privileged-information paradigm and provides a potentially reusable strategy for settings in which a costly, invasive, or sparsely available modality can guide the representation of a more scalable modality during training. In FORGE, this strategy combines cross-attention and FiLM-based fusion with a momentum-contrastive objective and representational-diversity regularization. The framework is also computationally tractable, training within a few minutes per epoch on a single accelerator and is released together with its phenotyping and evaluation tools to facilitate replication and extension.

Clinically, because the endophenotypes add discrimination against glaucoma beyond intraocular pressure, RNFL thickness and polygenic risk, they may have value for population-scale risk stratification once validated prospectively. More broadly, the same structure-function distillation strategy may be applicable wherever a sensitive functional measurement and a scalable structural modality coexist, including combinations of electrophysiology and imaging in other neurodegenerative disorders.

Several limitations constrain these conclusions. The genetic replication cohort was underpowered for variant-level genome-wide replication. External validation in UK Biobank was necessarily structural-only; consequently, the external analyses assessed the generalizability of the resulting OCT representation rather than the transfer of VF guidance itself. The external clinical-risk associations were also observational and should not be interpreted as evidence of causality. Training data were derived from a single tertiary-care center and were based on macular rather than peripapillary RNFL imaging. Finally, clustering granularity, although evaluated using two distinct algorithms, remains a modeling choice rather than a biologically predetermined quantity.

These limitations define the next steps. Expanding the genetic analysis to larger, multiancestry biobank cohorts, followed by statistical fine-mapping and colocalization with retinal expression and splicing quantitative-trait loci, will be necessary to move the candidate associations from nearest-gene proximity toward causal mechanism. Prospective and longitudinal evaluation will be required to determine whether the cross-sectional risk differences translate into clinically useful progression prediction and whether endophenotype-guided monitoring improves care. Methodologically, the privileged-information framework could be extended to additional functional modalities and to foundation-model backbones pretrained on large collections of unlabeled retinal images, potentially strengthening the representations before functional guidance is introduced. We release the code and phenotyping tools to facilitate these studies and enable independent evaluation of the resulting endophenotypes.

VF-guided entangled learning reframes mRNFL phenotyping as a structure-from-function problem: by treating the visual field as privileged information available only at training time, FORGE distils functional signal into the OCT representation and resolves clinically distinct endophenotypes with superior clustering coherence and divergent progression trajectories. Critically, these functionally informed phenotypes are not merely more separable, they are more heritable, recovering established glaucoma biology while surfacing novel loci near genes governing retinal development and axon guidance that structural-only definitions leave hidden. The learned null embedding is what makes this practical, collapsing the trained model to an OCT-only encoder that requires no perimetry at inference and generalizes to a population-scale cohort lacking functional data.

## Methods

The Institutional Review Board at Mass General Brigham approved this retrospective study, conducted in accordance with the Declaration of Helsinki. Given the retrospective design, the requirement for informed consent was waived.

### Development Dataset and Inclusion Criteria

MEE data (59,650 patients, 305,498 OCT scans; 76,767 patients, 441,006 VFs) were collected for training and validation. VF testing used standard automated perimetry (Humphrey Field Analyzer, 24-2 SITA-Standard) with quality criteria (FPR<30%, FL<20%, ≥3 VFs per patient). GL eyes were identified via PyGlaucoMetrics ^46^, and progression labels determined using vfprogression ^47^ ensemble criteria (MD slopes, Schell method ^48^, VFI), achieving 75% three-way agreement. Binary progression labels (0=not-worsening, 1=worsening) were assigned to 45,834 GL eyes with 227,259 labeled VFs (74% not-worsening, 26% worsening), providing auxiliary supervision during model training (Fig. 1A). OCT scans (Zeiss CIRRUS HD-OCT, 200×200 pattern, 6×6mm) underwent quality filtering (signal strength ≥7) and artifact correction (UNET partial convolution autoencoder ^49^), excluding images with >5% missing mRNFL values. Temporal matching (±6 months) transferred VF-derived labels to matched OCT scans. From 10,864 GL patients with 26,562 labeled VFs and 46,963 OCT scans, this process yielded 5,372 high-quality paired examinations from 1,845 patients (2,819 eyes). Model development used five-fold cross-validation with patient-level stratification; in each fold, 80% of patients (1,476 patients with 2,315 eyes and 4,834 scans) were used for training and validation and the remaining 20% (369 patients with 504 eyes and 538 scans) were held out for testing. All examinations and both eyes from a given patient were assigned exclusively to a single fold, preventing patient-level overlap and information leakage between training/validation and test sets (Fig. 9A).

**Fig. 9.**
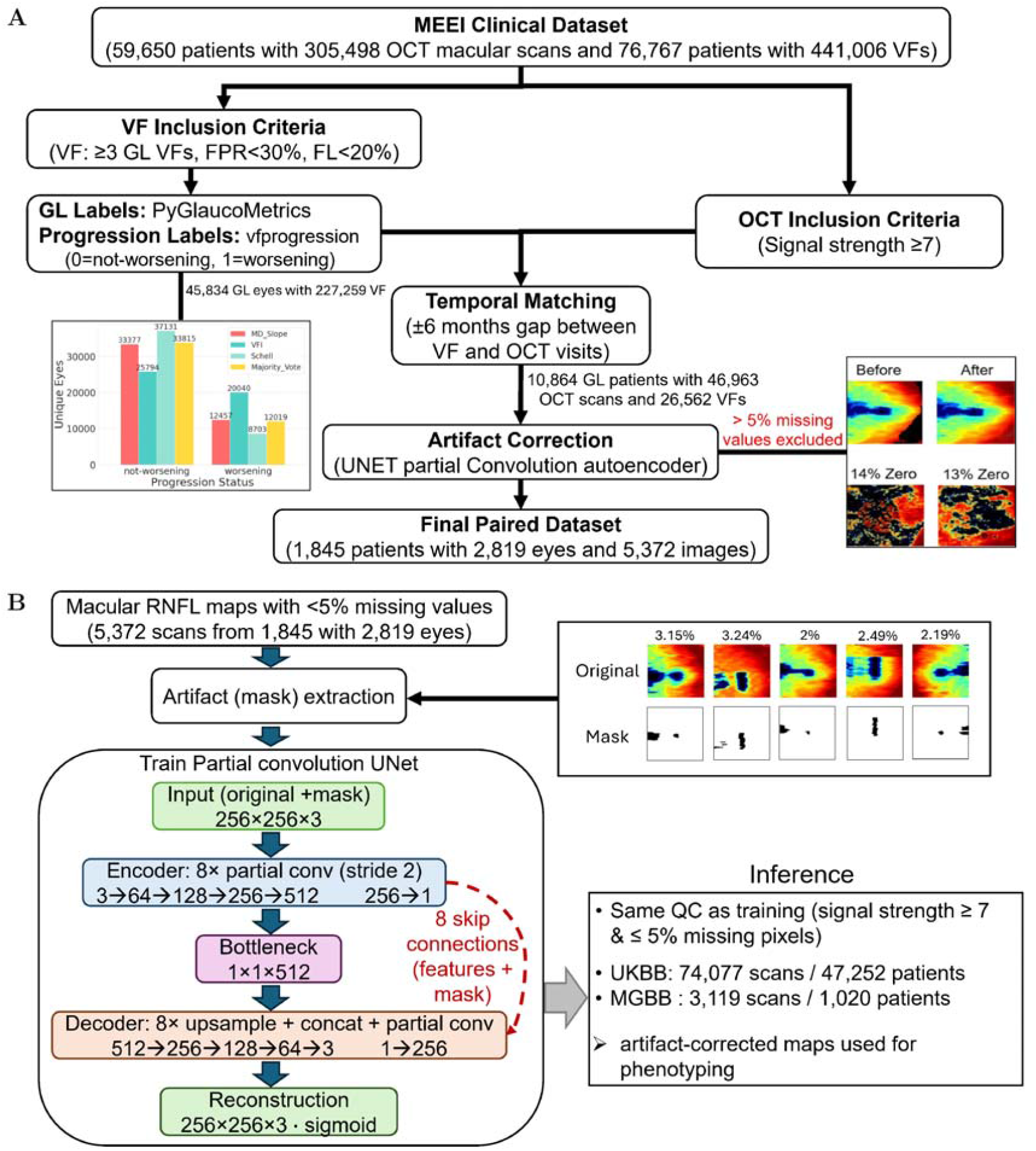
(A) MEE GL Dataset Curation Pipeline. Quality control applying inclusion criteria, temporal matching, and artifact correction. Examples show excluded OCT scans with poor image quality. (B) Artifact-correction pipeline. mRNFL maps with ≤5% missing pixels (5,372 scans; 1,845 patients; 2,819 eyes) were used to train a partial-convolution UNet with eight-stage encoder-decoder and skip connections in which each block renormalizes features by valid pixels and propagates the updated mask, to inpaint missing regions. Artifact masks were derived from the maps’ own zero-valued regions (right: examples labelled by missing-pixel fraction). The frozen model was then applied under identical quality control (signal strength ≥7, ≤5% missing) to UKBB (74,077 scans / 47,252 participants) and the MGBB (3,119 scans / 1,020 participants) for downstream phenotyping.

### Artifact Correction

Artifact correction was performed (Fig. 9B) using a previously validated partial convolution UNET adapted to mRNFL thickness maps ^49^. mRNFL thickness maps routinely contain missing (zero-valued) pixels caused by segmentation failure, vignetting, and vessel or blink shadows, which distort downstream phenotyping if left uncorrected. For every map we computed the fraction of missing pixels, and we retained only scans with ≤5% missing data (5,372 images from 1,845 patients with 2,819 eyes) for model development. The empirical missing regions of the retained maps were binarized into artifact masks (mask value 1 = observed, 0 = missing; Supplementary Fig. 18), providing a realistic, dataset-deriving corruption model rather than synthetic random holes.

Missing regions were reconstructed with a partial-convolution Unet ^50^, in which every convolution is renormalized by the valid pixels within its receptive field and the associated mask is updated layer by layer, so that reconstruction is conditioned only on observed data. The network was an eight-stage partial-convolution encoder-decoder with symmetric skip connections (encoder channels 3, 64, 128, 256, 512, using 7×7/5×5/3×3 partial convolutions with batch normalization and ReLU; decoder stages with nearest-neighbour upsampling, feature/mask concatenation, and LeakyReLU, ending in a three-channel sigmoid output). At each iteration an artifact mask was drawn at random and overlaid on a training image, and the model minimized the standard partial-convolution objective: L1 reconstruction on valid and hole regions (hole weighted ×6), VGG-16-based perceptual and style losses (x0.05 and x1), and a total-variation smoothness term (×0.1). Training used Adam (learning rate 2 × 10^−4^), batch size 4, 256x256 inputs, and frozen encoder batch-normalisation for small-batch stability, over 100 epochs; all loss components decreased smoothly and plateaued by 90 epochs, with total loss falling from 0.082 to 0.026 (Supplementary Fig. 19). The trained model was then applied to all MGBB and UKBB maps to inpaint missing or corrupted regions prior to phenotyping. The full partial-convolution UNet is shown in Supplementary Fig. 20.

### Evaluation Dataset

Two external cohorts were used to assess generalizability, the UKBB and the MGBB. Identical quality control to the MEE training set was applied to both (signal strength ≥ 7 and ≤ 5% missing pixels), retaining 74,077 high-quality mRNFL images from 47,252 UKBB participants and 3,119 images from 1,020 MGBB participants. All images were artifact-corrected with the pretrained partial-convolution UNet prior to phenotyping.

### Model Architecture and Training Protocols

The proposed framework (Fig. 1A) employs a shared ResNet50 backbone with separate mRNFL and VF encoders (256D) for momentum contrastive learning using VF as privileged supervision. The entangled fusion module integrates features through cross-attention (8 heads), Feature-wise Linear Modulation (FiLM) ^51^, 256D bottleneck, and momentum queue (K=4096), modulating mRNFL features via VF context:

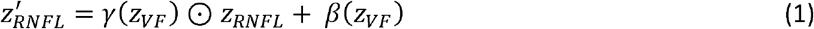

where *γ* and *β* are learned affine transformations from VF embeddings. The framework optimizes four loss components: MoCo aligns positive pairs while separating negatives:

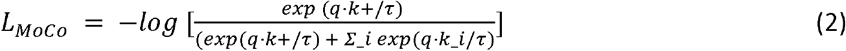

Cross-modal consistency enforces alignment between modalities:

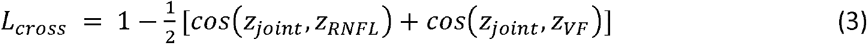

Diversity loss prevents representation collapse; *B* is batch size, *i* sample index.

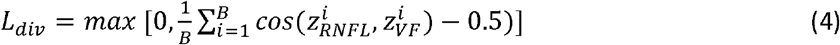

Progression classification provides auxiliary clinical supervision:

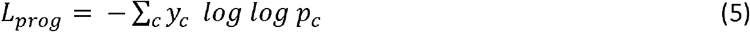

where *y*_*c*_ are progression labels and *p*_*c*_ are predicted probabilities. The total multi-objective loss combines these components:

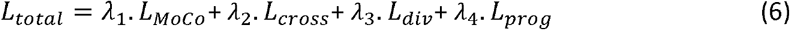

Loss weights (λ_1_=0.3, λ_2_=0.15, λ_3_=0.15, λ_4_=0.4) were optimized via Optuna ^52^ over 20 trials using validation clustering metrics, with ablation studies confirming each component’s contribution. All models were implemented in PyTorch 2.9 and trained on NVIDIA H100 GPUs (80GB), requiring ~2-3 minutes per epoch on the MEE dataset.

### Latent vector extraction and GWAS

External cohorts were phenotyped by pure feature transfer, without any retraining, refitting, or re-clustering on external data. The MEE-trained encoder was frozen and applied in inference mode to the artifact-corrected UKBB and MGBB maps, yielding a 256-dimensional multi-scale embedding per image from the fused layer-2, layer-3, and layer-4 representations of the ResNet-50 backbone. Because neither external cohort has paired perimetry, both were processed through the OCT-only path, in which the learned null embedding substitutes for the absent visual-field input. Embeddings were standardized using the scaler fitted on MEE, applied as a transform only, so that external data were mapped onto the internal feature distribution rather than re-normalized to their own, and then projected through the frozen β-VAE encoder, taking the deterministic posterior mean (*µ*) as the latent representation to avoid stochastic variation across inference runs. This yielded 15-dimensional (FCM) and 20-dimensional (GMM) latent vectors defined in the MEE latent space. Endophenotype assignment likewise used the frozen MEE cluster models: fuzzy c-means memberships were computed against the saved MEE centroids (m = 1.5), and Gaussian-mixture posteriors against the saved MEE components, so that cluster identity is directly comparable across all three cohorts.

Because participants could contribute multiple scans per eye, latent embeddings were reduced to one value per participant before association testing. For each patient-eye, only the most recent acquisition was retained, ranked by test date; for each participant, the most recent test date across both eyes was then identified and the left- and right-eye embeddings acquired on that date were averaged dimension-wise, yielding a single 15-dimensional (FCM) or 20-dimensional (GMM) phenotype vector per individual.

Association testing used the continuous latent axes as quantitative traits rather than cluster membership. Discretizing a continuous representation into cluster labels discards within-cluster variation and yields unbalanced categorical traits with substantially lower effective sample size per group, both of which reduce genome-wide power; this is the same rationale under which learned embeddings, rather than derived categorical labels, have been used as GWAS traits in other high-dimensional clinical and imaging modalities ^30,31^. All downstream analyses therefore use axis-level traits.

### Post Processing and Statistical Analysis

Latent vectors from *β*-VAE (Fig. 1B), obtained after freezing the encoder trained in Fig. 1A, were clustered using GMM with full/diagonal covariance and FCM with *m* ∈ {1.5, 2.0, 2.5, 3.0}, and grid search over k = 4-15. The selected latent dimensionality and clustering parameters (Fig. 1B) were fixed and applied to mRNFL-only embeddings for UKBB phenotyping (Fig. 1C). Clustering quality was evaluated using silhouette, Calinski-Harabasz, Davies-Bouldin, and diversity scores, with within-cluster similarity quantified using normalized cross-correlation (NCC) ^53^.

Statistical comparisons used paired t-tests for clustering metrics and Welch’s tests for NCC values (bootstrap n=1000, *P value* < 0.05), with MD slopes computed via mixed-effects models adjusted for age, severity, gender, race, and follow-up. Welch’s ANOVA was used for continuous variable comparisons across the three cohorts (age, baseline mRNFL, and IOP), Welch’s *t*-test for baseline MD (MEE vs MGBB), and Pearson *x*^2^ tests for categorical variables (sex and race). Welch’s corrections were applied because variances were unequal across cohorts. Given the large sample sizes, statistical significance was interpreted alongside absolute effect sizes.

### Phenotypic validation and incremental prediction

Phenotypic associations were tested in 44,477 genotyped European participants by logistic regression for binary outcomes and ordinary least squares for quantitative outcomes, adjusting for age, sex and ten genetic principal components. Because all 15 axes were non-normal by the Jarque-Bera test, primary analyses used rank-inverse-normal-transformed exposures with the Blom offset and likelihood-ratio tests; effects were expressed per 1 standard deviation. Sensitivity analyses used standardized and transformed scales, winsorization at |z| ≤ 4 and |z| ≤ 3, and exclusion of the extreme 0.1% and 1%.

Criterion validity included hospital-record POAG (ICD-10 H40.1, H40.8 or H40.9; 325 cases), any glaucoma including self-report (849 cases), cataract, age-related macular degeneration, diabetic retinopathy and retinal detachment. Convergent validity used Goldmann-correlated IOP, corneal resistance factor, and cup-disc ratio. Discriminant validity used osteoarthritis, asthma, gallstones, inguinal hernia, breast cancer, myocardial infarction and depression. All disease definitions were prevalent rather than incident, and case and denominator count for every outcome, together with the per-subcode counts behind the two glaucoma definitions, are given in Supplementary Table 21.

Incremental discrimination was assessed by five-fold cross-validated AUROC using ridge-penalized logistic regression with nested cross-validation for the penalty and five outer seeds. Four cumulative models included age and sex; the same plus Goldmann-correlated intraocular pressure and mean macular retinal nerve fibre layer thickness; the same plus a glaucoma polygenic score; and the same plus the 15 endophenotypes. All models used 34,411 participants with both clinical measurements. The polygenic score was constructed with PRS-CS ^54^ from FinnGen R12 H7_GLAUCOMA_POAG (10,832 cases, 473,757 controls) ^55^. The two outcomes were hospital-record POAG, with 235 cases after analysis-set restriction, and any glaucoma, with 626 cases. Pairwise model comparisons used paired DeLong tests ^56^ on pooled out-of-fold predictions, with a cross-seed variance included in the confidence intervals. As a control for the number of added predictors, model 4 was repeated with the 15 endophenotypes replaced by permuted (label-shuffled) copies, preserving their marginal distributions and intercorrelation structure while removing any association with outcome, through the identical nested-CV, ridge-penalized pipeline.

### Genotype data and quality control

The UK Biobank is a prospective cohort of approximately 500,000 participants aged 40 to 69 recruited across the United Kingdom between 2006 and 2010 ^57^. Participants were genotyped on the Applied Biosystems UK BiLEVE Axiom Array or UK Biobank Axiom Array. The Mass General Brigham Biobank (MGBB) contains more than 200,000 consented patient samples linked to electronic health records ^58^. MGBB participants were genotyped on three versions of the Illumina biobank array. The cohorts share no participants and were genotyped, imputed and quality-controlled independently using different arrays and reference panels.

Sample quality control was performed with PLINK 1.9 and PLINK 2.0 ^59^ in 488,395 genotyped UK Biobank participants. Withdrawn participants and those absent from the imputed release or without quality-control data were removed, together with 1,930 participants of ambiguous sex, 978 ancestry-specific heterozygosity outliers and 35,113 participants with Pi-hat > 0.1875. Genetic ancestry was inferred from LD-pruned markers (r2 < 0.2 in 200 kb windows, MAF > 0.05) using the first five principal components and k-nearest-neighbors classification against 1000 Genomes Project Phase 3 labels ^60^. Of 47,252 participants, 44,477 had genotypes and were assigned European genetic ancestry. Requiring non-missing spherical-equivalent refractive error gave the primary GWAS set of N = 39,381.

Directly genotyped variants (GRCh37) were retained at minor allele frequency > 0.01, minor allele count > 100, missingness < 0.1 and Hardy-Weinberg equilibrium P > 1 × 10^−15^, leaving 629,669 variants for ridge regression. Imputed autosomal variants were filtered at posterior probability > 0.9, INFO ≥ 0.6, ancestry-specific minor allele frequency ≥ 0.001 and Hardy-Weinberg equilibrium by the mid-P exact test, followed by minor allele frequency > 0.01. Chromosome X and the pseudoautosomal region were excluded, leaving 9,329,555 autosomal variants. A per-variant call-rate threshold of N ≥ 35,443 (≥ 90% of 39,381) retained 9,050,596 variants per trait.

### Genome-wide association analysis

Association testing used REGENIE v4.1.2 ^61^ two-step whole-genome ridge regression. Step 1 fitted per-trait polygenic predictors on the 629,669 array variants (block size 1,000); step 2 used leave-one-chromosome-out predictors as offsets for single-variant testing across the imputed panel (block size 400, minimum minor allele counts 20). All 15 endophenotypes were analyzed as quantitative traits and rank-inverse-normal transformed within REGENIE (-qt --apply-rint). Models included the first ten genetic principal components, age and spherical-equivalent refractive error as quantitative covariates, and sex and genotyping array as categorical covariates. Age was taken from the recruitment assessment (field 21022), because age at imaging was unavailable. Effects were conditional on refractive error. Inflation was assessed by the median-χ2 genomic control factor and the univariate LD score regression intercept.

Independent signals were defined by LD clumping in PLINK 2.0 using in-sample linkage disequilibrium from the 39,381 participants, with r2 < 0.1, a ±500 kb window, index P < 5 × 10^−^8. Per-endophenotype index variants were pooled across traits and re-clumped under the same parameters; leads within 500 kb were collapsed into physical regions. Leads were annotated to the nearest protein-coding gene and nearest gene of any biotype using GENCODE v47 (GRCh38) ^62^ and classified as coding, untranslated region, non-coding-transcript exon, intronic or intergenic.

Novelty was assessed against published POAG and IOP genome-wide significant associations and the previous OCT-only endophenotype meta-analysis [10.21203/rs.3.rs-8311177/v1]. The POAG and IOP published associations were extracted from three major POAG studies ^27,63^ and from the NHGRI-EBI GWAS Catalog associations for glaucoma (MONDO:0005041 with child traits) and intraocular pressure (EFO:0004695), downloaded 2025-04-05; the studies contributing to the two Catalog downloads, with their PubMed identifiers and attributed variant counts, are listed in Supplementary Table 22. Matches were classified hierarchically as LEAD, an exact chromosome-position match; PROXY, an LD proxy within ±1 Mb; PROXIMITY, a reference variant within ±500 kb without an exact or proxy match; or no match. Loci without a match in any source were considered novel. Proxy analyses used r2 ≥ 0.2 and r2 ≥ 0.8 with UK Biobank-derived and MGBB European LD panels.

### Multiple-testing thresholds

The effective number of endophenotypes was estimated by the Li-Ji method as M_eff_ = 5. Genome-wide significance was defined as P < 5 × 10^−8^, with 5 × 10^−8^/5 = 1.0 × 10^−8^ reported as the M_eff_ tier. Gene-based, transcriptome-wide and splicing-wide analyses used 0.05/(n_features_ × 15) as the primary threshold and 0.05/(n_features_ × 5) as the effective-dimensionality threshold. Phenotypic validation used Benjamini-Hochberg FDR across 15 endophenotypes within each outcome and a study-wide threshold of 0.05/(Meff × 3 outcome families) = 3.33 × 10^−3^. Colocalization was Bayesian and did not use a significance threshold.

### Heritability and genetic architecture

SNP heritability was estimated by LD score regression (LDSC) ^64^ and GCTA-GREML (v1.94.1) ^65^. LDSC used rsID-bearing summary statistics munged against the HapMap 3 ^66^, precomputed European LD scores and approximately 1.16 million variants per trait; heritability was reported on the observed scale. GCTA-GREML used a genetic relationship matrix from the 629,669 array variants. Removing one individual from each pair with GRM relatedness > 0.05 left N = 37,749, with rank-inverse-normal-transformed phenotypes and the GWAS covariates.

Cross-trait LDSC estimated genetic correlations among all 105 endophenotype pairs using a Bonferroni threshold of 0.05/105 = 4.8 × 10^−4^. Effective dimensionality was calculated from the phenotypic and 15 × 15 genetic correlation matrices using Li-Ji. Agreement between phenotypic and genetic correlations was summarised by the orientation-invariant uncentred correlation and sign concordance.

To assess visual-field guidance, the representation was refitted without it, producing 20 latent axes. Both representations were analyzed in the same 39,381 participants with the same genotype panel, REGENIE v4.1.2 model, transformation, covariates, 90% call-rate filter and locus-definition pipeline. Because the axes had no one-to-one correspondence, distributions were compared by their medians using the Mann-Whitney U test. Heritability analyses were repeated after restricting the ablation to its 15 axes with the highest heritability Z. Ablation loci were also evaluated with the same novelty and cell-type-enrichment procedures.

### Replication in the Mass General Brigham Biobank

MGBB array variants were excluded at minor allele frequency < 0.05, missingness > 0.05, batch-bias P < 5 × 10^−5^, Hardy-Weinberg equilibrium P < 1 × 10^−10^ and for palindromic alleles. Cleaned arrays were phased with SHAPEIT4, imputed to the TOPMed ^67^ r^2^ panel through the Michigan Imputation Server ^68^, union-merged, and retained at R2 > 0.6 and post-imputation missingness < 0.05. European ancestry was assigned by random forest classification after projection onto HGDP and 1000 Genomes principal components ^69^. Of 64,145 genotyped participants, 53,852 were assigned European ancestry and 53,846 were present in the released genotype file.

The endophenotypes were measured in 714 European MGBB participants; 711 remained after exclusion at KING kinship > 0.0884 ^70^. GRCh38 hard-called variants were analysed at MAF > 0.01. Each lead variant was tested only against the endophenotypes for which it reached *P* < 5 × 10^−8^ in discovery, leaving the 105 pairs over 53 variants in 26 independent 1 Mb loci. The association test was performed with PLINK 2 --glm, adjusting for sex, age, spherical equivalent, missing spherical equivalent and ten principal components. Replication was summarized by sign concordance, discovery-replication effect correlation and an effect-size ratio estimated by Deming regression with λ = mean (SE^2^_MGBB_)/mean (SE^2^_discovery_) = 46.8. Inference used locus-clustered permutations and locus-block bootstrap intervals. Clinical associations were tested in n = 435 participants with ascertainable POAG status (280 cases, 155 controls) by logistic regression with Benjamini-Hochberg FDR across 15 axes, adjusting for age, sex, and 10 within-sample PCs. Per-pair replication power was estimated under a normal approximation using the noncentrality parameter |β_discovery_|/SE_MGBB_, if the replication effect equaled the discovery estimate. Power was evaluated for one- and two-sided tests at nominal α=0.05 and Bonferroni-corrected α=0.05/105=4.8×10^−4^.

### Statistical fine mapping

Each of the 41 physical regions was fine-mapped in its strongest trait with SuSiE-RSS (susieR v0.14.2) ^71^, using regional z scores and an external LD matrix to estimate 95% credible sets and posterior inclusion probabilities. Models used L = 10, 95% coverage and fixed residual variance. LD matrices were made positive semi-definite by R ← (R + λI)/(1 + λ), with median λ = 0.026. UK Biobank-derived (N = 337,000) and MGBB European (N = 53,846) LD panels were evaluated; the UK Biobank panel was used for the reported analysis.

### Gene-based association

MAGMA v1.10 ^72^ was applied to call-rate-filtered summary statistics. Variants were assigned to NCBI build 37.3 genes using a 35 kb upstream and 10 kb downstream window, and gene P values were estimated by the SNP-wise mean model with the 1000 Genomes European panel (503 participants) and N = 39,381. A total of 18,417 genes were tested per trait. Predicted expression and splicing were tested with S-PrediXcan ^73^ and S-MultiXcan ^74^ from MetaXcan (metax v0.7.5) using PredictDB GTEx v8 ^75^ MASHR models on GRCh38 (DOI 10.5281/zenodo.3518299). Eighteen tissues were selected, including tibial nerve as an optic-nerve proxy and all 13 brain regions. The 540 S-PrediXcan analyses (15 traits × 18 tissues × 2 layers) were combined across tissues into 30 S-MultiXcan gene-level scans.

### Colocalization

Colocalization between the strongest association signal in each region and cis-e/sQTL was tested with coloc (v5.2.3) ^76^ and susieR (v0.14.2) ^77^. The GTEx analysis compared the regional SuSiE-RSS credible-set log-Bayes-factor matrix with precomputed SuSiE matrices from the eQTL Catalogue (study QTS000015) ^78^, allowing multiple causal variants per region. The retina analysis used coloc.abf() with EyeGEx cis-eQTL ^79^ and assumed one causal variant because no LD-matched retina panel was available. Regions with < 70% EyeGEx variant matching were excluded. Priors were p1 = p2 = 1 × 10−4 and p12 = 5 × 10^−6^. Calls required PP4 ≥ 0.8, PP4/(PP3 + PP4) ≥ 0.9 and ≥ 500 shared variants, followed by visual validation using locus-comparison plots.

### Summary-data-based tests of cis expression and splicing

SMR (v1.03) ^80^ with the HEIDI test and a 1000 Genomes European LD reference was used to distinguish shared association from variants in tight linkage. The instruments per endophenotype was restricted to the 41 fine-mapped physical regions extended by 3 Mb on each side, retaining 887,457 variants per trait. EyeGEx cis-eQTL provided 15 genes × 15 endophenotypes = 225 retina tests. GTEx V8 BESD data for twelve tissues, namely aorta and tibial artery, cerebellar hemisphere, cerebellum, cortex, hippocampus, hypothalamus and putamen, cultured fibroblasts, tibial nerve as the optic-nerve proxy, subcutaneous adipose and whole blood yielded 634 genes and 36,825 expression tests. The corresponding cis-sQTL data produced 2,002 introns by tissue pairs across 196 genes and 30,030 splicing tests (Supplementary Table 23). Genes were called when they passed the SMR test and HEIDI P > 0.05. Retina expression, GTEx expression and GTEx splicing were corrected as separate families at 2.2 × 10^−4^, 1.36 × 10^−6^ and 1.67 × 10^−6^, respectively.

### Locus-based cell-type enrichment with ECLIPSER

ECLIPSER was applied using the Segrè laboratory Snakemake implementation ^81^ to 3,647 variants reaching P < 1 × 10−6 across the 15 endophenotypes; the pipeline retained variants at P ≤ 5 × 10^−8^ before clumping. This identified 32 loci, of which 31 had mapped genes. Gene mapping used GTEx v8 cis-eQTL and cis-sQTL supplemented by Open Targets Genetics locus-to-gene assignments ^82^. Enrichment was assessed in retina, macula, optic nerve head and anterior segment references as the Bayesian Fisher ratio of leading-edge loci among trait and null GWAS loci, with 95% credible intervals and one-sided P values. Benjamini-Hochberg correction was reported within each reference.

## Data Availability

All data produced in the present study are available upon reasonable request to the authors.

## Data Availability

The data that support the findings of this study can be obtained from the corresponding author upon reasonable request. Summary statistics of the UKBB GWAS will be shared publicly upon publication.

## Code Availability

The codes used in this study are publicly available in the first author’s GitHub page at: https://github.com/Mousamoradi/.

## Acknowledgment

This research was funded by the National Institutes of Health (R01 EY036518, R01 EY030575, R01 EY038687, R01 EY036222, and P30 EY003790), and Research to Prevent Blindness.

## Author Contribution

M.M. data collection, conceptualization, methodology, algorithm design, data curation, software (code development), formal analysis, validation, visualization, writing original draft, review & editing. L.C. GWAS analysis, visualization, review & editing. Y.Z. GWAS runs, GWAS analysis, visualization, review & editing. S.S. data collection, review & editing. M.W. and T.E. resources, funding acquisition. M.E. conceptualization, visualization, review & editing. A.S. validation, visualization, review & editing. J. W. validation, visualization, review & editing. N.Z. conceptualization, validation, visualization, review & editing, project administration, resources, funding acquisition.

